# Association between Use of Qingfei Paidu Tang and Mortality in Hospitalized Patients with COVID-19: A national retrospective registry study

**DOI:** 10.1101/2020.12.23.20248444

**Authors:** Lihua Zhang, Xin Zheng, Xueke Bai, Qing Wang, Bowang Chen, Haibo Wang, Jiapeng Lu, Shuang Hu, Xiaoyan Zhang, Haibo Zhang, Jiamin Liu, Ying Shi, Zhiye Zhou, Lanxia Gan, Xi Li, Jing Li

## Abstract

**Background:** Qingfei Paidu Tang (QPT), a formula of traditional Chinese medicine, which was suggested to be able to ease symptoms in patients with Coronavirus Disease 2019 (COVID-19), has been recommended by clinical guidelines and widely used to treat COVID-19 in China. However, whether it decreases mortality remains unknown.

**Purpose:** We aimed to explore the association between QPT use and in-hospital mortality among patients hospitalized for COVID-19.

**Study design:** A retrospective study based on a real-world database was conducted.

**Methods:** We identified patients consecutively hospitalized with COVID-19 in 15 hospitals from a national retrospective registry in China, from January through May 2020. Data on patients’ characteristics, treatments, and outcomes were extracted from the electronic medical records. The association of QPT use with mortality was evaluated using Cox proportional hazards models based on propensity score analysis.

**Results:** Of the 8939 patients included, 28.7% received QPT. The crude mortality was 1.2% (95% confidence interval [CI] 0.8% to 1.7%) among the patients receiving QPT and 4.8% (95% CI 4.3% to 5.3%) among those not receiving QPT. After adjustment for patient characteristics and concomitant treatments, QPT use was associated with a relative reduction of 50% in in-hospital mortality (hazard ratio, 0.50; 95% CI, 0.37 to 0.66 *P* <0.001). This association was consistent across subgroups by sex and age. Meanwhile, the incidence of acute liver injury (8.9% [95% CI, 7.8% to 10.1%]vs. 9.9% [95% CI, 9.2% to 10.7%]; odds ratio, 0.96 [95% CI, 0.81% to 1.14%], *P* =0.658) and acute kidney injury (1.6% [95% CI, 1.2% to 2.2%] vs. 3.0% [95% CI, 2.6% to 3.5%]; odds ratio, 0.85 [95% CI, 0.62 to 1.17], *P* =0.318) was comparable between patients receiving QPT and those not receiving QPT. The major study limitations included that the study was an observational study based on real-world data rather than a randomized control trial, and the quality of data could be affected by the accuracy and completeness of medical records.

**Conclusions:** QPT was associated with a substantially lower risk of in-hospital mortality, without extra risk of acute liver injury or acute kidney injury among patients hospitalized with COVID-19.

## Introduction

Coronavirus Disease 2019 (COVID-19), caused by a novel severe acute respiratory syndrome coronavirus type 2 (SARS-CoV-2), has posed a huge threat to global health as the largest pandemic in a century. Nearly 50 million people worldwide have been infected, of whom over 1.2 million died by middle November2020.^1^ The pandemic is still evolving, effective treatments against COVID-19 are therefore urgently needed to reduce the mortality of COVID-19.

Qingfei Paidu Tang (QPT), a traditional Chinese medicine, was formulated on the basis of one of the classics of traditional Chinese medicine, Treatise on Febrile and Miscellaneous Diseases (*Shang Han Zabing Lun*).^2^ It is a compound prescription containing four traditional Chinese medicine prescriptions, each of which has been widely applied as therapy of common cold, fever, influenza, and other virus infection.^3-7^ Basic research also found that QPT possessed properties such as antivirus, ^8,9^ anti-inflammation, ^8-13^ and immune regulation,^8,11-13^ which might be beneficial for patients with COVID-19. Moreover, several small observational studies in China have suggested its potential effectiveness in relieving symptom (i.e., fever and cough) and preventing disease progression in patients with COVID-19.^14-17^ Therefore, QPT has been recommended in the Chinese guidelines for the treatment of Coronavirus Disease 2019 (COVID-19) since early February 2020 and widely used in China.^18^ However, it is unknown whether it could reduce the mortality of COVID-19.

Accordingly, using the data from a national retrospective registry, we sought to evaluate the effectiveness and safety of QPT in COVID-19. Specifically, we hypothesized that QPT use would be associated with a lower risk of in-hospital mortality in patients with COVID-19, and tested it using propensity score analysis. We also assessed whether there was an association of QPT with the incidence of acute liver injury and acute renal injury during hospitalization.

## Methods

### Data Sources

In a government-mandated national registry, hospitalizations for COVID-19 in all the designated hospitals across China were registered retrospectively. Information relating to patient characteristics, treatments, and outcomes, in the electronic medical records (EMR), were required to be submitted to a system deployed by the National Health Commission of China, in forms of either structured database for the front page, or unstructured text for the progress notes, lab test results, and physician’ s orders. By the date of May 6^th^ 2020, over 40 thousand COVID-19 cases from more than five hundred hospitals have been included.

### Ethical approval

The Ethics Committee at the the National Center for Cardiovascular Diseases (NCCD)/Fuwai Hospital ethics committee approved this study and the Ethics Committee at the First Affiliated Hospital, Sun Yat-sen University approved the current analysis. Informed consent of individual patients was waived.

### Study cohort

Among all the designated hospitals providing inpatient care for COVID-19 in the national registry, we excluded hospitals that were ineligible for data extraction or analysis for the following two reasons. First, the number of patients hospitalized with COVID-19 was less than 100. Second, the number of patients receiving QPT in the hospitals was less than 50. In the end, 15 hospitals were included in our study, all of which were located in Hubei province.

Among eligible hospitals, we included all patients aged ≥18 years discharged between January and May, 2020 with a confirmed diagnosis of COVID-19. We identified these patients, according to International Classification of Diseases, Clinical Modification codes revision 10 (U07.100, U07.100.00x, U07.100.00×001, U07.100.00×002, U07.100.00×003), when available, or through principal diagnosis terms noted at discharge. We excluded patients who were transferred out, since the records of their hospitalizations were truncated. Patients who died or were discharged within 24 hours of admission were also excluded from the analysis, because the testing and treatments for them were likely to be influenced due to the short length of hospital stay.

### Data Extraction

For each patient, the demographic characteristics (age and sex), prior medical histories/comorbidities (hypertension, diabetes, coronary heart disease, stroke, chronic kidney diseases, chronic obstructive pulmonary disease, and cancer), clinical status at admission (critical or not), and in-hospital death was obtained from the front-page database or progress notes. The vital signs (heart rate, blood pressure, and respiratory rate) at admission were extracted from the progress notes. The in-hospital medications (QPT, Arbidol, Ribavirin, Oseltamivir, Ganciclovir, Lopinavir, Lianhuaqingwen, Xuebijing, Diammonium Glycyrrhizinate, Methylprednisolone, Dexamethasone, and Interferon) were extracted from the physician orders, progress notes, and nurse records. The in-hospital acute liver injury and acute kidney injury were identified based on the front-page database, progress notes, and lab test results.

We searched predefined keywords in unstructured text of the submitted medical records using Python software (version 3.6) and MYSQL software (version 8.0), in order to extract the data. Particularly, research clinicians randomly selected and reviewed 5% of the medical records in the hospitals with QPT use rate under 20%, to ensure the exhaustion of synonyms of this medication and completeness of data extraction. Furthermore, to ensure the data accuracy, research clinicians adjudicated the prior medical history/comorbidities based on the progress note.

### Treatment and outcome measures

As the treatment of interest in our analysis, QPT use was defined as receiving this medication for no less than three days during the hospitalization, according to the Chinese diagnosis and treatment protocol for COVID-19 (Trial Version 6) (i.e., one formula a day, three formulas were defined as a course of treatment).^18^ Correspondently, the study cohort was categorized into two treatment groups – patients receiving QPT and those not receiving QPT. Meanwhile, we also explored the effectiveness and safety of QPT between patients who ever received QPT during hospitalization and those who did not.

The outcome measure of effectiveness was in-hospital mortality. The outcome measure of safety included acute liver injury and acute kidney injury during hospitalization. Acute liver injury was defined as documented acute liver injury, acute liver renal insufficiency, acute liver failure, hepatic encephalopathy, or hepatic coma, then adjudicated based on the elevation in aspartate aminotransferase, alanine aminotransferase, or total bilirubin. Acute renal injury was defined as documented acute renal failure, acute renal injury, or acute renal insufficiency, then adjudicated based on the elevation in serum creatinine.

### Statistical analysis

We described participant characteristics, treatments, and outcomes, with frequencies and percentages for categorical variables, while means ± standard deviations or median with interquartile range (IQR) for continuous variables. The difference between groups was estimated by standard mean difference (SMD), and absolute values less than 0.1 was considered small differences.^19^

We conducted a statistical power analysis in advance, based on the projected sample size of this retrospective registry. Assuming the in-hospital mortality rate was 4% in patients not receiving QPT, a total sample size of 9000 can achieve a statistical power of 80% at a 2-sided 0.05 significance level to detect a hazard ratio of 0.7 or below, for the treatment with a 30% or greater prevalence.

We used inverse probability treatment weighting (IPTW) based on probability of receiving treatment to make the characteristics between the two treatment groups comparable. The probability of receiving QPT was estimated by multilevel logistic regression that adjusted for baseline characteristics including demographics, comorbidities, and prior histories extracted in previous referred (Table S1), with hospital as random effect.

To assess the effectiveness of QPT, we obtained hazard ratios (HR) between treatment groups with developing frailty proportional hazards models on in-hospital death, accounted hospital as random effect, adjusted for other in-hospital medications, and weighted with inverse probability of QPT use. We then plotted Kaplan-Meier curve in patients receiving and those not receiving QPT. To assess the safety of QPT, we obtained odds ratios (OR) with the multilevel logistic regression on acute liver injury and acute renal injury, which handled random effect, adjustment, and weight, using the similar approaches described earlier. We also added interaction items to explore the heterogeneity of effectiveness across subgroups by age (<60, 60-69, or ≥ 70 years), sex (male or female), and prior medical history/comorbidities (with any or without). In each subgroup, we recalculated inverse probability and reweighting separately, as aforementioned.

We conducted two sensitivity analysis. First, we matched propensity score between patients receiving and not receiving QPT using the nearest-neighbour method, to create two groups with similar characteristics and sample size. Second, we added the propensity score as covariate in the frailty model without weighting, to account for the difference between treatment groups.

In the submitted medical records, small proportions of blood pressure (1.7%), heart rate (0.1%), and respiratory rate (0.2%) were missing. Assuming that these data were missing at random, we applied a multiple imputation method based on Markov Chain Monte Carlo by PROC MI procedure in SAS to impute missing value.^20^

Two-tailed P values were reported with P<0.05 considered to indicate statistical significance. All statistical analyses were performed with SAS software, version 9.4 (SAS Institute, Cary, NC).

## Results

### Study Participants

There were 9115 patients with COVID-19 admitted to the 15 designated hospitals in this study, with the numbers of cases in each included hospital ranging from 140 to 1856. After excluding 96 patients with age <18 years, 66 patients transferred out, and 14 patients with the length of stay less than 24 hours, 8939 eligible cases were included in the analysis (Figure 1). Of them, the average age was 55.9±15.6 years, and 53.4% (4771) were women. 4.4% (390) patients were at critical state at admission, while 33.7% (3016) had hypertension, and 15.2% (1357) had diabetes.

**Figure 1.**
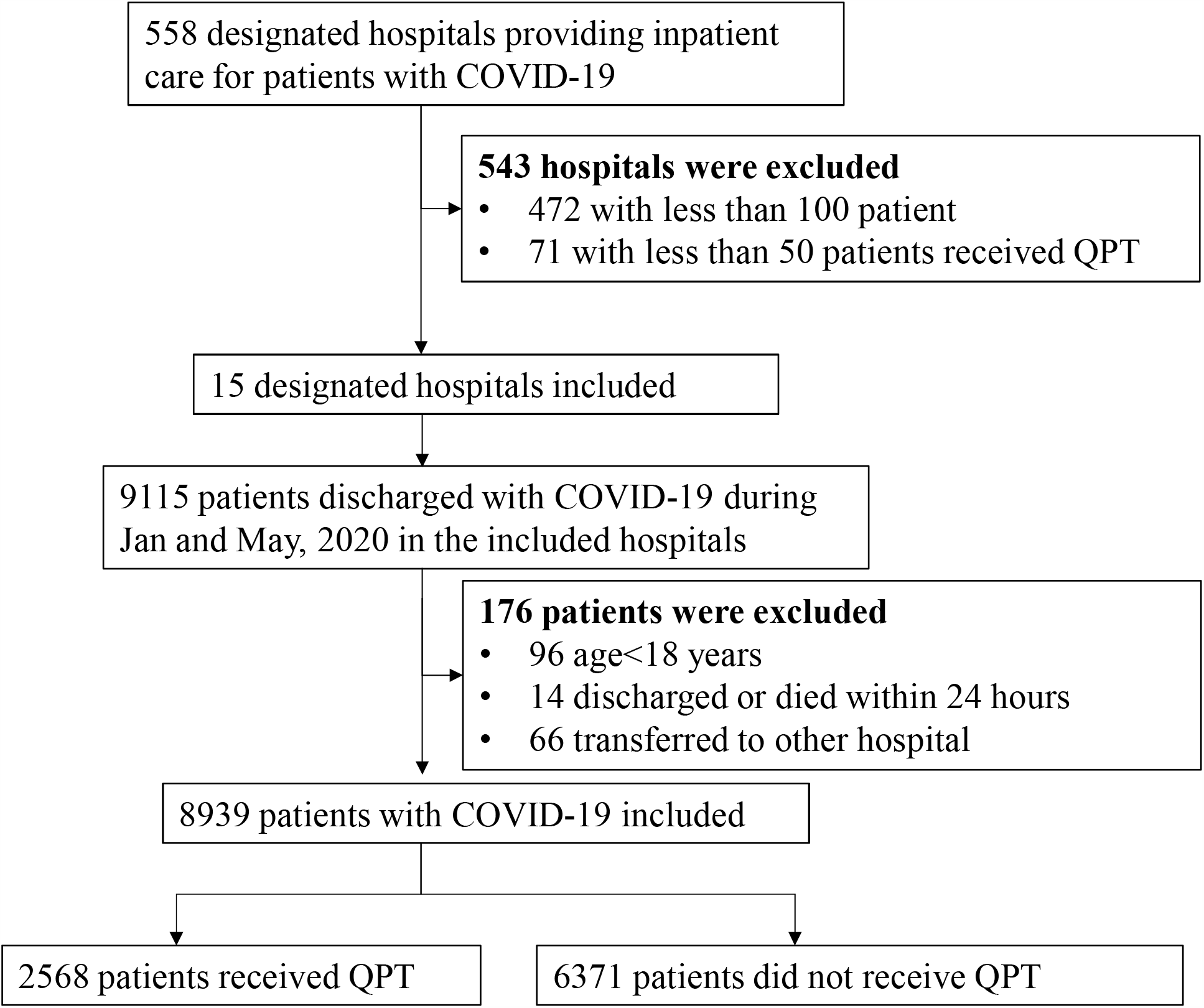
Flowchart of the study cohort. COVID-19, coronavirus disease 2019; QPT, Qingfei Paidu Tang

Of these patients, 2833 (31.7%) ever received QPT during hospitalization, with a median treatment duration of 6 (4 to 9) days. Half of the QPT users received the first formula within 5 days after hospitalization. The timing of QPT use after hospitalization was shown in Figure S1.

In the study cohort, 2568 patients (28.7%) received QPT for no less than 3 days and 6371 (71.3%) did not. The patient characteristics of the two treatment groups were provided in Table 1. Unweighted comparisons showed that patients who received QPT were younger (SMD>0.1). After adjustment for inverse probability of treatment weighting, all covariates were well balanced (i.e., standardized mean differences were <0.1). The distributions of inverse probability score weights of treatment groups were shown separately in Figure S2.

**Table 1.**
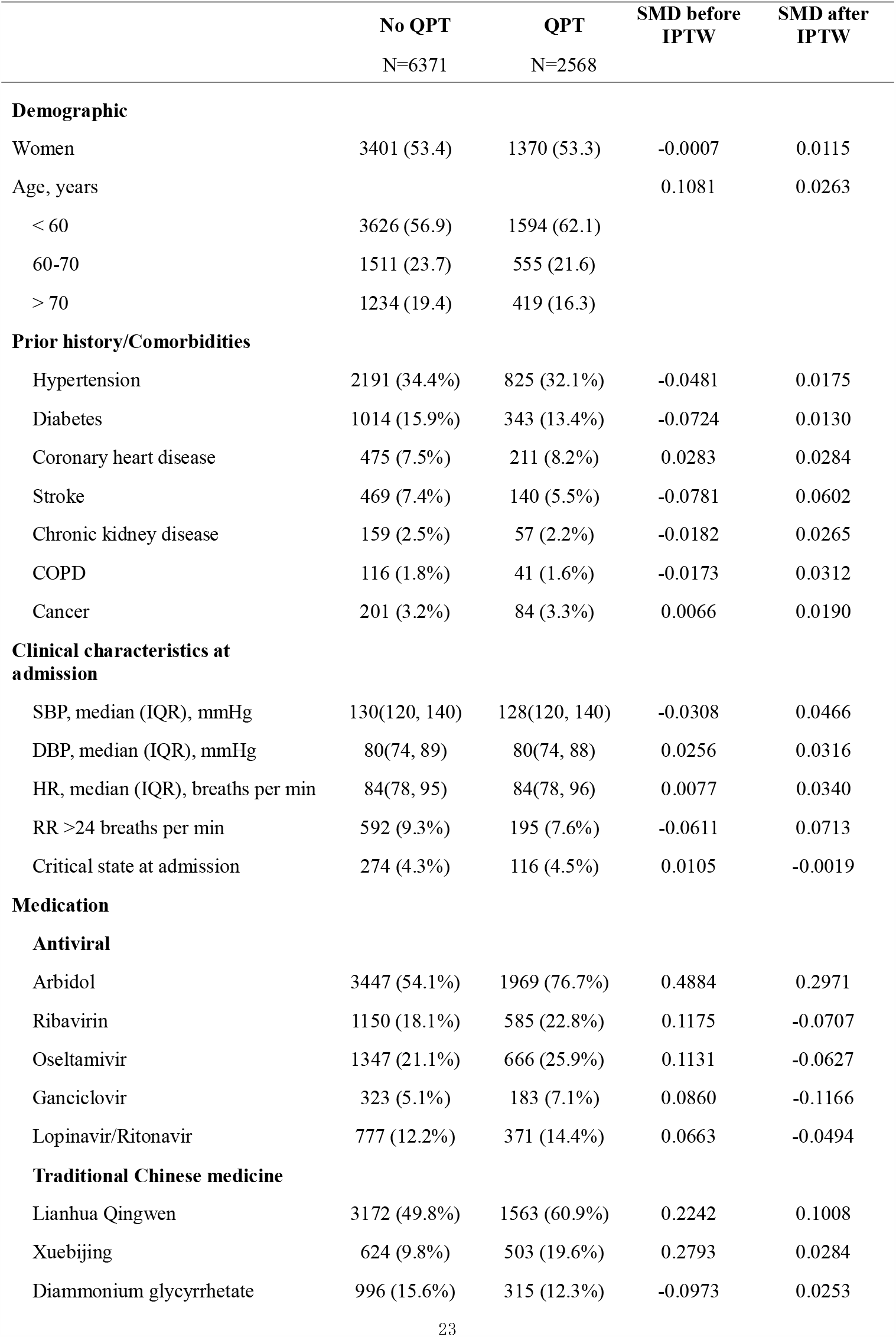
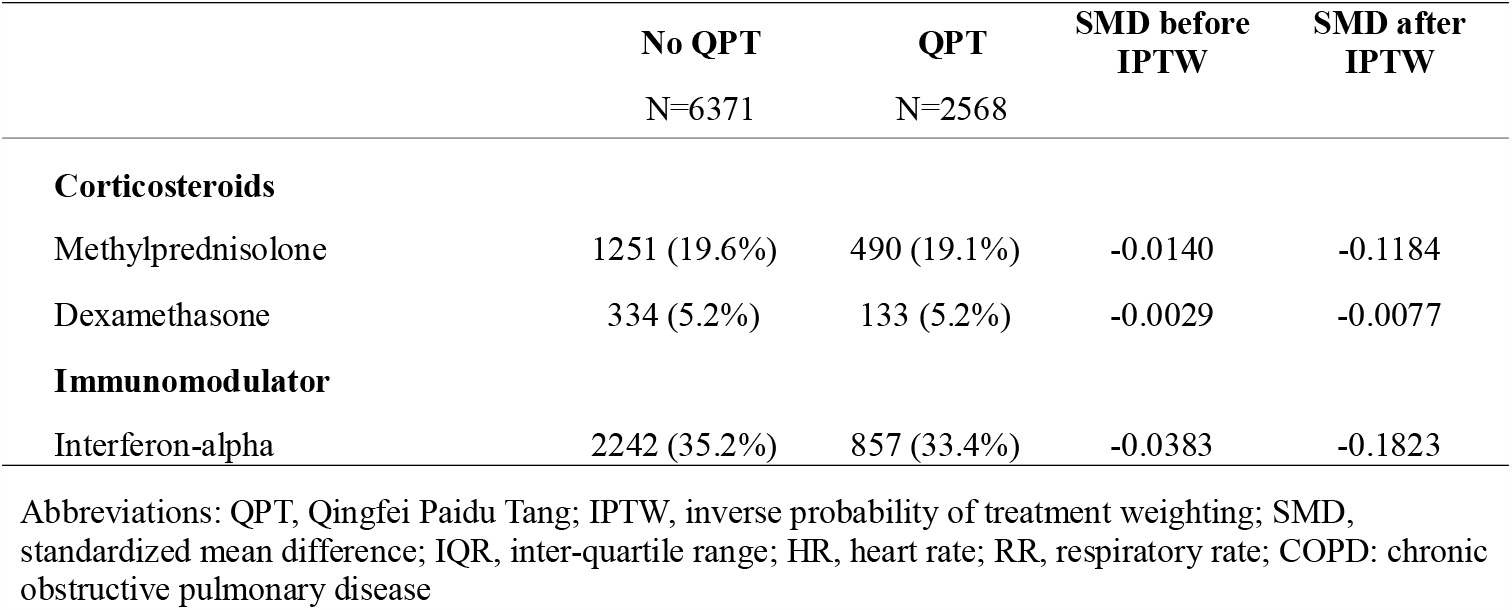
Baseline characteristics of patients by Qingfei Paidu Tang use.

|  | No QPT<br>N=6371 | QPT<br>N=2568 | SMD before<br>IPTW | SMD after<br>IPTW |
| --- | --- | --- | --- | --- |
| **Demographic** |  |  |  |  |
| Women | 3401 (53.4) | 1370 (53.3) | -0.0007 | 0.0115 |
| Age, years |  |  | 0.1081 | 0.0263 |
| < 60 | 3626 (56.9) | 1594 (62.1) |  |  |
| 60-70 | 1511 (23.7) | 555 (21.6) |  |  |
| > 70 | 1234 (19.4) | 419 (16.3) |  |  |
| **Prior history/Comorbidities** |  |  |  |  |
| Hypertension | 2191 (34.4%) | 825 (32.1%) | -0.0481 | 0.0175 |
| Diabetes | 1014 (15.9%) | 343 (13.4%) | -0.0724 | 0.0130 |
| Coronary heart disease | 475 (7.5%) | 211 (8.2%) | 0.0283 | 0.0284 |
| Stroke | 469 (7.4%) | 140 (5.5%) | -0.0781 | 0.0602 |
| Chronic kidney disease | 159 (2.5%) | 57 (2.2%) | -0.0182 | 0.0265 |
| COPD | 116 (1.8%) | 41 (1.6%) | -0.0173 | 0.0312 |
| Cancer | 201 (3.2%) | 84 (3.3%) | 0.0066 | 0.0190 |
| **Clinical characteristics at admission** |  |  |  |  |
| SBP, median (IQR), mmHg | 130(120, 140) | 128(120, 140) | -0.0308 | 0.0466 |
| DBP, median (IQR), mmHg | 80(74, 89) | 80(74, 88) | 0.0256 | 0.0316 |
| HR, median (IQR), breaths per min | 84(78, 95) | 84(78, 96) | 0.0077 | 0.0340 |
| RR >24 breaths per min | 592 (9.3%) | 195 (7.6%) | -0.0611 | 0.0713 |
| Critical state at admission | 274 (4.3%) | 116 (4.5%) | 0.0105 | -0.0019 |
| **Medication** |  |  |  |  |
| **Antiviral** |  |  |  |  |
| Arbidol | 3447 (54.1%) | 1969 (76.7%) | 0.4884 | 0.2971 |
| Ribavirin | 1150 (18.1%) | 585 (22.8%) | 0.1175 | -0.0707 |
| Oseltamivir | 1347 (21.1%) | 666 (25.9%) | 0.1131 | -0.0627 |
| Ganciclovir | 323 (5.1%) | 183 (7.1%) | 0.0860 | -0.1166 |
| Lopinavir/Ritonavir | 777 (12.2%) | 371 (14.4%) | 0.0663 | -0.0494 |
| **Traditional Chinese medicine** |  |  |  |  |
| Lianhua Qingwen | 3172 (49.8%) | 1563 (60.9%) | 0.2242 | 0.1008 |
| Xuebijing | 624 (9.8%) | 503 (19.6%) | 0.2793 | 0.0284 |
| Diammonium glycyrrhetate | 996 (15.6%) | 315 (12.3%) | -0.0973 | 0.0253 |
| **Corticosteroids** |  |  |  |  |
| Methylprednisolone | 1251 (19.6%) | 490 (19.1%) | -0.0140 | -0.1184 |
| Dexamethasone | 334 (5.2%) | 133 (5.2%) | -0.0029 | -0.0077 |
| **Immunomodulator** |  |  |  |  |
| Interferon-alpha | 2242 (35.2%) | 857 (33.4%) | -0.0383 | -0.1823 |

Abbreviations: QPT, Qingfei Paidu Tang; IPTW, inverse probability of treatment weighting; SMD, standardized mean difference; IQR, inter-quartile range; HR, heart rate; RR, respiratory rate; COPD: chronic obstructive pulmonary disease

### Outcomes

During hospitalization with a median length of stay of 15 (9 to 21) days, 334 (3.7%) patients died. The crude mortality was 1.2% (95% confidence interval [CI], 0.8% to 1.7%) among patients who received QPT and 4.8% (95% CI, 4.3% to 5.3%) among patients who did not (Figure 2). In the unadjusted analysis, patients who received QPT were less likely to die than patients who did not receive QPT (hazard ratio, 0.17; 95% CI, 0.11% to 0.26%, *P*<0.001). In the Cox model with inverse propensity score weighting, all covariates in the Cox model were shown in Table S2. QPT use was associated with a lower mortality risk (adjusted hazard ratio, 0.50; 95% CI, 0.37 to 0.66, *P*<0.001).

**Figure 2.**
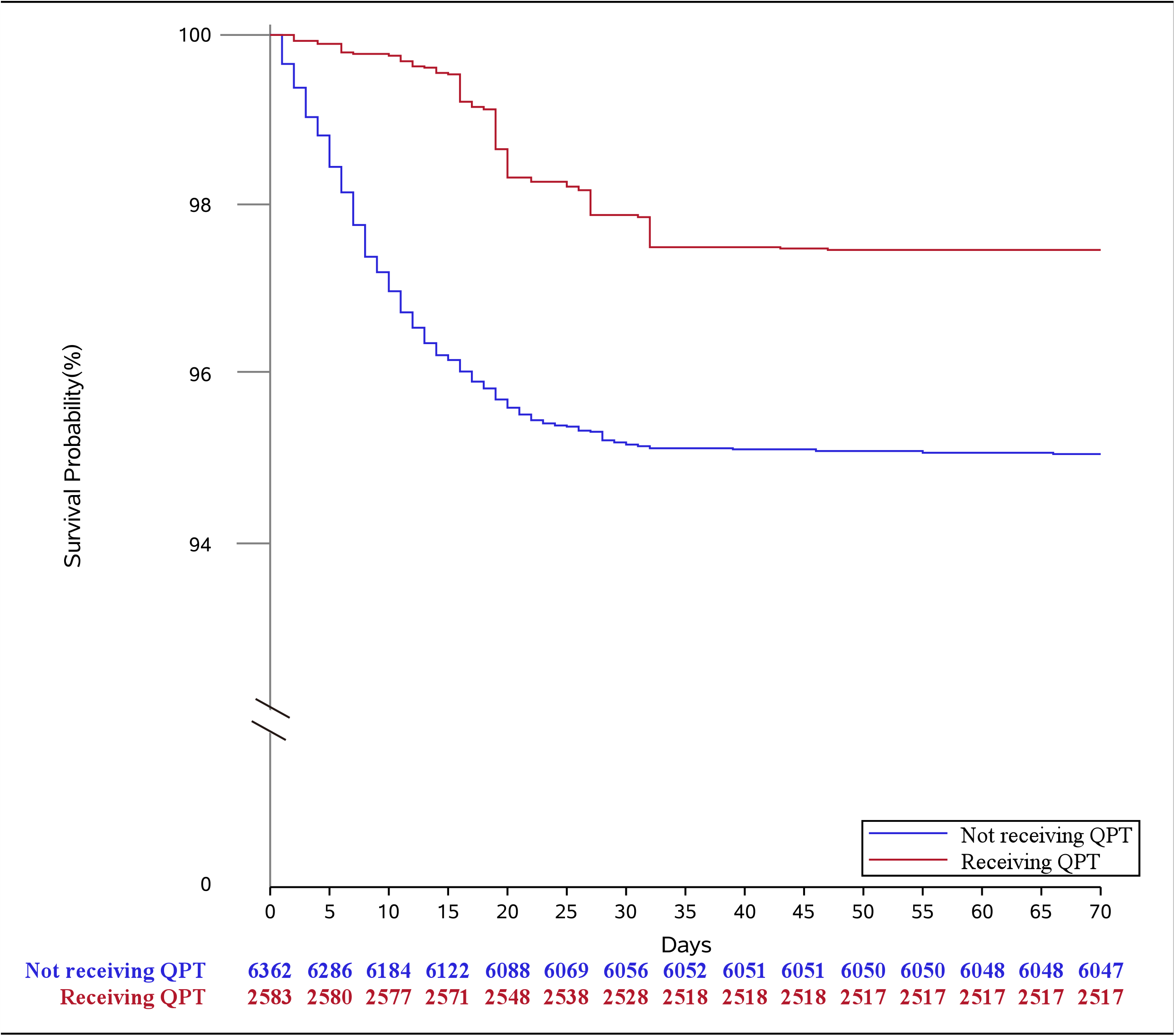
Kaplan–Meier survival curves for in-hospital mortality in inverse probability treatment weighting analysis. QPT, Qingfei Paidu Tang

In terms of sex and age, no significant differences were observed among their subgroups in the associations between QPT treatment and in-hospital mortality (all *P* for interaction>0.05). Although significant heterogeneity in associations between QPT treatment and in-hospital mortality were detected between subgroups by prior medical history/comorbidities status (*P* for interaction=0.020), the significantly lower mortality risk for patient receiving QPT was observed in both these subgroups (Figure 3).

**Figure 3.**
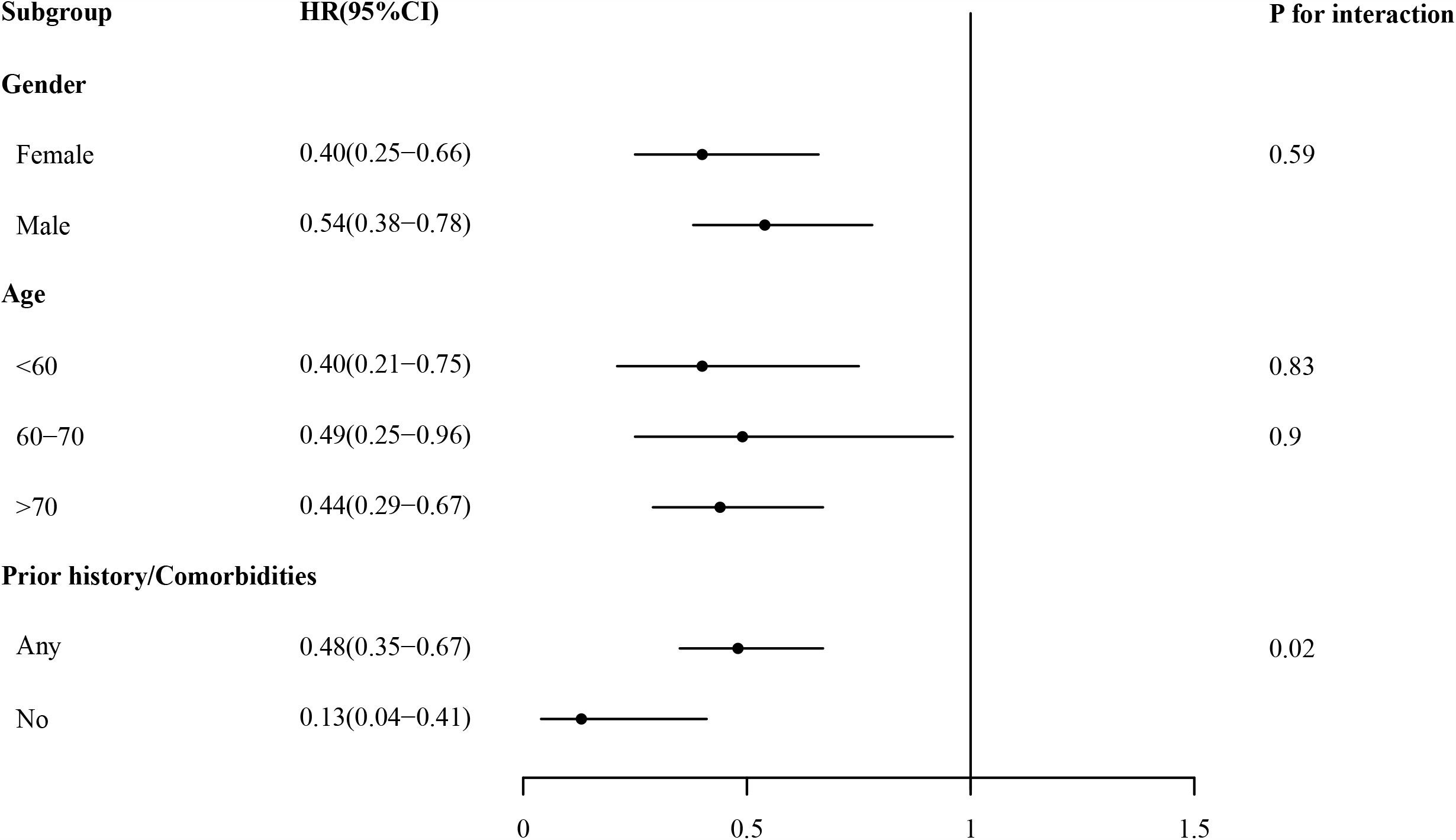
Hazard ratios of in-hospital mortality across subgroups in inverse probability treatment weighting analysis. HR, hazard ratio; 95% CI, 95% confidence interval.

Regarding the safety of QPT, patients who received QPT had a comparable incidence of acute hepatic injury (crude rate, 8.9% [95% CI, 7.8% to 10.1%] vs 9.9% [95% CI, 9.2% to 10.7%]; adjusted OR, 0.96 [95% CI, 0.81 to 1.14], *P* =0.658) and acute kidney injury (crude rate, 1.6% [95% CI, 1.2% to 2.2%] vs. 3.0% [95% CI, 2.6% to 3.5%]; adjusted OR, 0.85 [95% CI, 0.62 to 1.17], *P* =0.318), in comparison with those who did not.

Furthermore, we also conducted the analysis of the effectiveness and safety of QPT between patients who ever received QPT during hospitalization and those who did not, and found similar results with those mentioned above (Table S3-4).

### Sensitivity Analyses

In addition to the IPTW analysis, we matched 3492 patients based on their propensity score (1746 patients receiving QPT and 1746 patients not receiving QPT). The two groups were well-balanced in characteristics and concomitant treatments (Table S5, Figure S3). The risk of mortality in patients who received QPT was significantly lower than in those who did not receive QPT (1.1% [95% CI, 0.7% to 1.7%] vs 2.7% [95% CI, 2.0% to 3.6%], HR, 0.42; 95%CI, 0.24 to 0.74; *P*= 0.002) (Table 2 and Figure S4). In the meantime, patients receiving QPT had a comparable incidence of acute kidney injury (1.1% [95% CI, 0.7% to 1.8%] vs. 1.9% [95% CI, 1.3% to 2.6%]; OR, 0.74 [95% CI, 0.40 to 1.35], *P* =0.327) compared with the patients who did not, but a lower risk of acute liver injury (5.4% [95% CI, 4.4% to 6.5%]vs. 8.1% [95% CI, 6.9% to 9.5%]; OR, 0.72 [95% CI, 0.54 to 0.96], *P* =0.025).

**Table 2.**
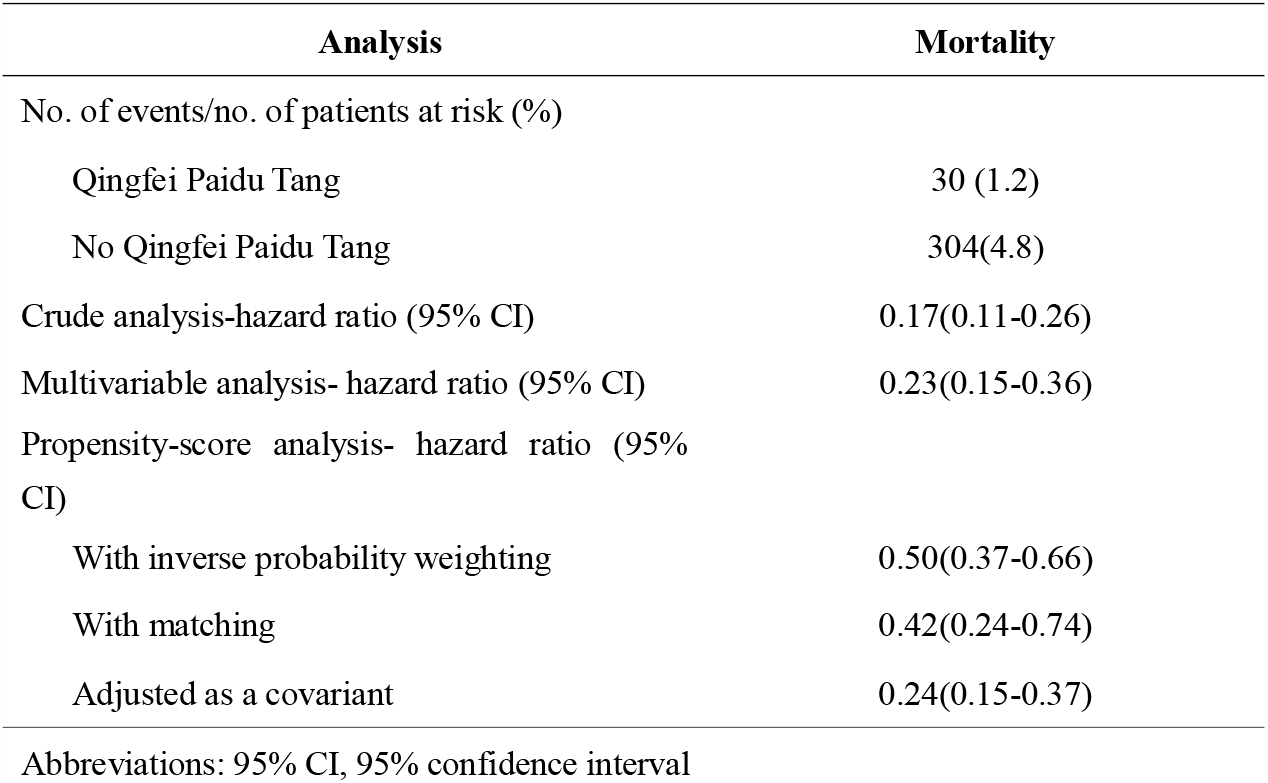
Associations between Qingfei Paidu Tang use and mortality in the crude analysis, multivariable analysis, and propensity-score analyses.

| Analysis | Mortality |
| --- | --- |
| No. of events/no. of patients at risk (%) |  |
| Qingfei Paidu Tang | 30 (1.2) |
| No Qingfei Paidu Tang | 304(4.8) |
| Crude analysis-hazard ratio (95% CI) | 0.17(0.11-0.26) |
| Multivariable analysis- hazard ratio (95% CI) | 0.23(0.15-0.36) |
| Propensity-score analysis- hazard ratio (95% CI) |  |
| With inverse probability weighting | 0.50(0.37-0.66) |
| With matching | 0.42(0.24-0.74) |
| Adjusted as a covariant | 0.24(0.15-0.37) |

Abbreviations: 95% CI, 95% confidence interval

We also included the propensity score as an additional covariate in the models, in which patients who received QPT had a significantly lower risk of mortality than those who did not receive QPT (adjusted HR, 0.24 95% CI, 0.15 to 0.37; *P*<0.001). Meanwhile, patients receiving QPT had comparable incidence of acute liver injury (OR, 0.93 [95% CI, 0.76 to 1.14], *P* =0.497) and acute kidney injury (OR, 0.74 [95% CI, 0.50 to 1.10], *P* =0.133) compared with the patients not receiving QPT.

## DISCUSSION

In this analysis based on a national registry of hospitalized patients with COVID-19, we first demonstrated that QPT use was associated with halving the risk of in-hospital mortality, without significant increase in risk of adverse effects, such as acute liver injury or acute kidney injury. Our findings have provided new evidence and insights regarding the treatment of COVID-19.

Our study has extended the literature on the effectiveness of QPT for patients with COVID-19. First, this is the first study assessing the association between the QPT use and in-hospital mortality that is considered the most important and objective outcome metrics, rather than surrogate indicators widely used before. Second, in comparison with prior studies in China about QPT for COVID-19 treatment,^14-17^ our study has involved an over ninety-time larger sample size that ensured sufficient statistical power even for subgroup analysis. Third, using various propensity score approaches, we established control groups to enable appropriate comparisons in both effectiveness and safety of QPT. Forth, this national registry included consecutive patients from multiple Chinese hospitals, which represented the use and effectiveness of QPT in real-world practice.

The effects of QPT on decreasing mortality of COVID-19 observed in our study are supported by the mechanisms shown in prior experimental studies, which included antivirus,^8,9^ anti-inflammation,^8-13^ immune regulation,^8,11-13^ regulating metabolism, ^9,12^ anti-platelet aggregation,^10^ and organ protection.^11,13^ QPT was composed of four traditional Chinese medicine prescriptions, which were shown to be separately effective in antivirus,^3,5^ anti-inflammatory,^7^ or immuno-modulating.^6^ QPT has multiple components acting on the multiple pathways. Some studies employed molecular network and network pharmacology to analyse the ingredients of QPT, and found that the key active ingredients, including quercetin, luteolin, kaempferol, naringenin, and isorhamnetin, could alleviate excessive immune responses, by regulating the function of cytokines related pathways, such as tumour necrosis factor signalling pathways and mitogen-activated protein kinases signalling pathways.^11-13^ Further research is needed to fully investigate the underlying mechanism of the effect of QPT.

In this study, we did not observe the elevated risk of acute liver injury or acute kidney injury among patients receiving QPT. This is consistent with the previous observational studies.^14-17^ Moreover, our findings are particularly reassuring given the complexity in comorbidities (such as hypertension, diabetes and chronic kidney disease) and concomitant treatments (such as antivirals, corticosteroids and immunomodulators) observed in our cohort. Nevertheless, long-term safety related to QPT still needs to be verified in future studies.

This study has provided valuable evidence and prospects for the treatment of COVID-19. Currently, there are globally nearly 7.5 million active cases that need treatments.^1^ However, there is no evidence about any medication that could decrease mortality in COVID-19 except for dexamethasone, which has been proved to be able to reduce the 28-day mortality in those who received mechanical ventilation or oxygen alone.^21,22^ To the best of our knowledge, this is the first study implying that QPT could reduce the mortality risk of patients with COVID-19. Our findings were consistent across subgroups, and robust regardless of analytic methods. It is encouraging that the use of QPT can probably prevent tens of thousands of deaths, if our findings are further confirmed and applied globally.

## Limitations

The results of our study should be interpreted in the context of several limitations. First, due to the nature of observational study, we cannot exclude the influence of residual confounders. However, after the IPTW, patients who received QPT had higher rates of co-morbidities which was positively related to mortality risk, compared with those who did not received QPT. Thus, the effectiveness we observed tended to be conservative. Second, our study was based on real-world data and the quality of data could be affected by the accuracy and completeness of medical records. Therefore, we only included the highly reliable variables on patient characteristics, treatments, and outcomes in the analysis. Third, our study merely collected in-hospital outcomes, therefore, we could not evaluate the long-term effectiveness and safety.

Finally, all the patients in our study were from China, and the beneficial effects of QPT in other racially diverse populations still await further validation.

## Conclusion

Among the patients hospitalized for COVID-19, the use of QPT was associated with halving the risk of mortality, without raising the risk of acute liver injury or acute kidney injury. Further validation with randomized controlled trials is needed to support the use of QPT worldwide for COVID-19.

## Data Availability

The data sharing needs to be approved by national registry, which is under the supervision of National Health commission. However, on site data audit is allowed under current regulation.

## Declarations

### Ethics approval

The Ethics Committee at the First Affiliated Hospital, Sun Yat-sen University approved the current analysis. Informed consent of individual patients was waived.

### Consent for publication

Not applicable

### Authors’ contributions

JL and XL conceived of the project and take responsibility for all aspects of it. JL and XL designed the study. LHZ and XZ wrote the first draft of the manuscript, with further contributions from JPL, XKB, HBZ, JML, BWC, QW, XYZ, HS, HBW, YS, ZYZ and LXG. XKB and SH did the statistical analysis. YZW, ZYZ, ESS, WQ, BWC, YS and XYZ collected, extracted, processed, and cleaned the data. All authors have read and approved the submission of this review, which neither has been published on any other peer-review platforms, nor is being considered for publication elsewhere, in whole or in part, in any language.

## Acknowledgements

We appreciate the Bureau of Medical Administration and Medical Service Supervision, National Health Commission of China, for the approval and support on collecting data. We appreciate all the COVID-19 designated hospitals for submitting medical records, and China Standard Medical Information Research Center for the support in collecting and processing data. We appreciate the team in National Center for Cardiovascular Diseases for their multiple contributions in data cleaning and manuscript coordinating.

## Abbreviations

QPT: Qingfei Paidu Tang
COVID-19: Coronavirus Disease 2019
IQR: interquartile range
SMD: standard mean difference
IPTW: inverse probability treatment weighting
HR: hazard ratios
OR: odds ratios
CI: confidence interval

