## Supplementary Appendix for "Association between Use of Qingfei Paidu Tang and Mortality in Hospitalized Patients with COVID-19: A national retrospective registry study"

**Table of content**

**Page 2 Figure S1.** Histogram of time (in days) to first formula of Qingfei Paidu Tang for those patients that received Qingfei Paidu Tang before discharge

**Page 3 Figure S2.** Distribution of inverse probability score weights in patients receiving Qingfei Paidu Tang and patients who did not receive Qingfei Paidu Tang.

**Page 4 Figure S3**. Distribution of the estimated propensity score for receiving Qingfei Paidu Tang, among patients who did and did not receive Qingfei Paidu Tang

**Page 5 Figure S4**. Kaplan-Meier curve for in hospital mortality by Qingfei Paidu Tang treatment status with propensity score matching

**Page 6 Table S1.** Odds ratios (95% CIs) of receiving Qingfei Paidu Tang treatment for all variables included in the propensity score model

**Page 7 Table S2.** Hazard ratios (95% CIs) of all variables included in the inverse probability weighted Cox model

**Page 8-9 Table S3.** Baseline characteristics of patients by Qingfei Paidu Tang use

**Page 10 Table S4.** Associations between Qingfei Paidu Tang use and mortality, acute hepatic injury, acute kidney injury

**Page 11-12 Table S5**. Baseline characteristics of patients by Qingfei Paidu Tang use status using propensity score matching

**Figure S1.** Histogram of time (in days) to first formula of Qingfei Paidu Tang for those patients that received Qingfei Paidu Tang before discharge

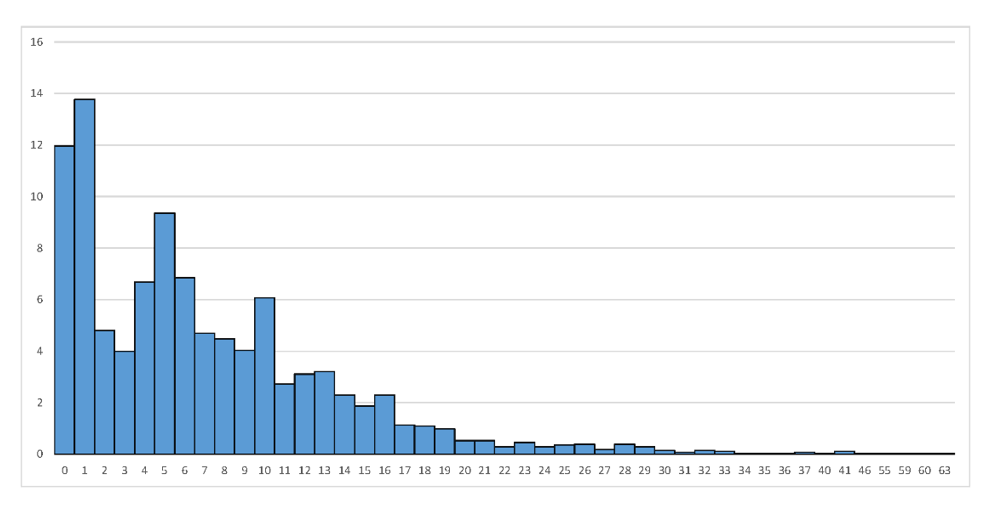

**Figure S2.** Distribution of inverse probability score weights in patients receiving Qingfei Paidu Tang and patients who did not receive Qingfei Paidu Tang

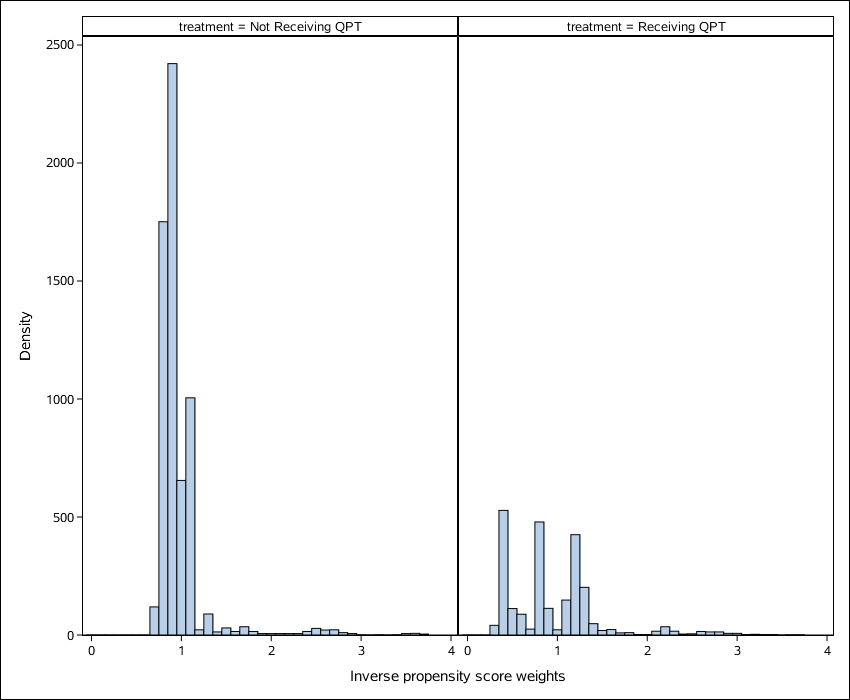

The weights were truncated by resetting the value of weights greater (lower) than percentile 99 (1) to the value of percentile 99 (1)

**Figure S3.** Distribution of the estimated propensity score for receiving Qingfei Paidu Tang, among patients who did and did not receive Qingfei Paidu Tang

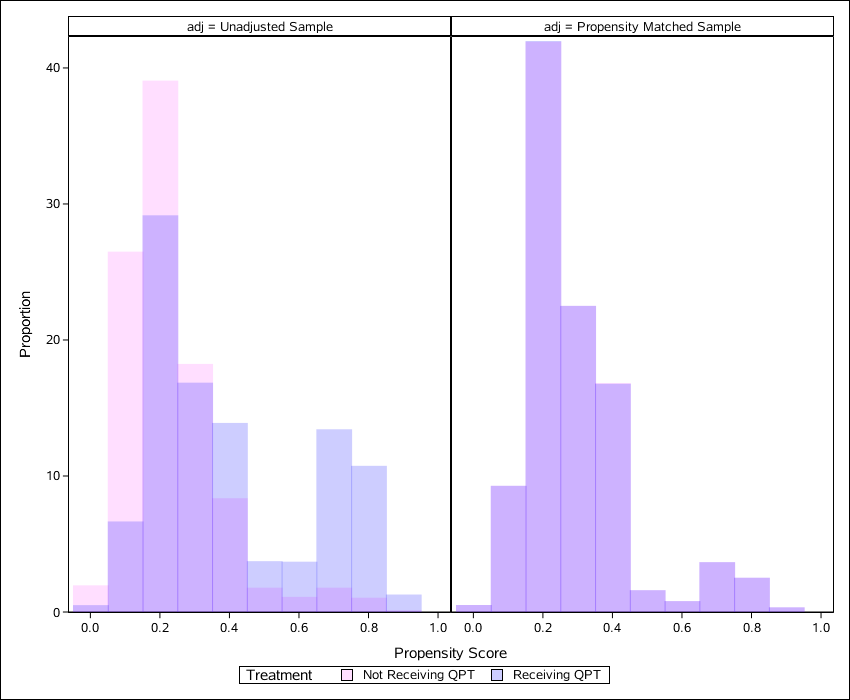

QPT indicates Qingfei Paidu Tang

**Figure S4.** Kaplan-Meier curve for in hospital mortality by Qingfei Paidu Tang treatment status with propensity score matching

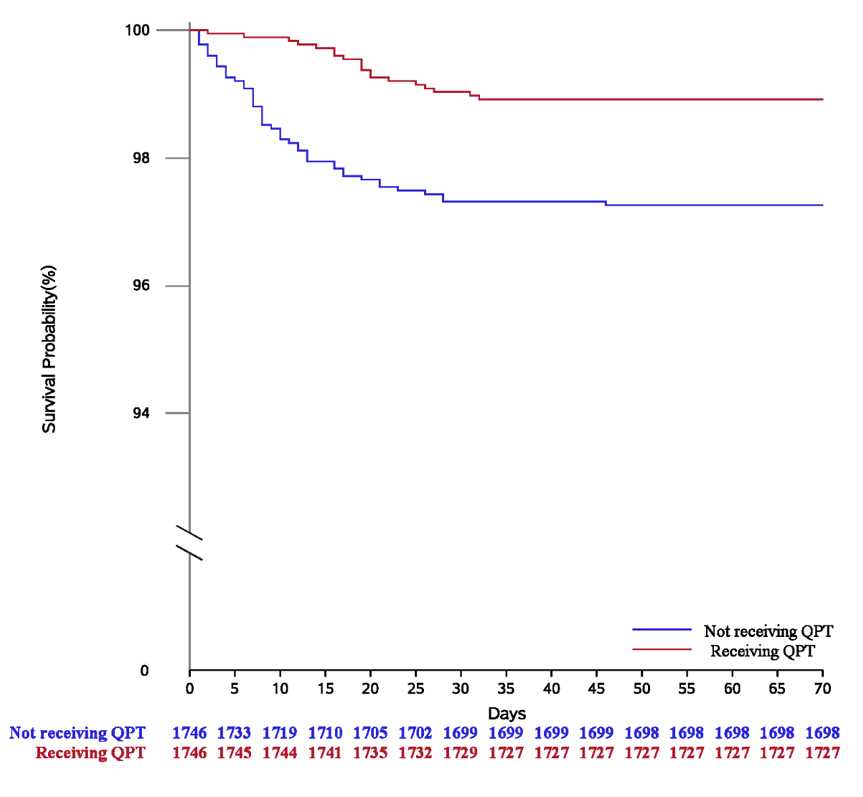

QPT indicates Qingfei Paidu Tang

**Table S1. Odds ratios (95% confidence intervals) of receiving Qingfei Paidu Tang treatment for all variables included in the propensity score model**

|  | **Odds ratios**  **(95% confidence interval)** | ***P* value** |
| --- | --- | --- |
| **Demographic** |  |  |
| Women | 0.96(0.87-1.06) | 0.426 |
| Age, years |  |  |
| <60 | 1.00 |  |
| 60-70 | 0.96 (0.84-1.10) | 0.578 |
| >70 | 0.94 (0.81-1.10) | 0.439 |
| **Clinical characteristics at admission** |  |  |
| Systolic blood pressure, mmHg | 1.00(1.00-1.00) | 0.784 |
| Diastolic blood pressure, mmHg | 1.00(0.99-1.00) | 0.893 |
| Heart rate, beats per min | 1.00(1.00-1.00) | 0.389 |
| Respiratory rate >24 breaths per min | 0.92(0.76-1.12) | 0.403 |
| Critical state at admission | 0.73(0.54-0.97) | 0.030 |
| **Prior history/Comorbidities** |  |  |
| Hypertension | 1.02(0.88-1.16) | 0.723 |
| Diabetes | 0.90(0.77-1.05) | 0.169 |
| Coronary heart disease | 1.10(0.89-1.36) | 0.367 |
| Stroke | 0.69(0.55-0.87) | 0.002 |
| Chronic kidney disease | 0.70(0.49-1.01) | 0.054 |
| Chronic obstructive pulmonary disease | 0.92(0.62-1.37) | 0.686 |
| Cancer | 0.80(0.59-1.09) | 0.151 |

**Table S2. Hazard ratios (95% CIs) of all variables included in the inverse probability weighted Cox model**

| **Variables** | **Hazard ratios**  **(95% confidence interval)** | ***P* value** |
| --- | --- | --- |
| Qingfei Paidu Tang | 0.50(0.37-0.66) | <0.001 |
| Arbidol | 0.68(0.54-0.86) | <0.001 |
| Ribavirin | 1.00(0.76-1.30) | 0.976 |
| Oseltamivir | 0.77(0.61-0.97) | 0.030 |
| Ganciclovir | 0.87(0.61-1.23) | 0.436 |
| Lopinavir/Ritonavir | 1.35(1.02-1.79) | 0.036 |
| Lianhua Qingwen | 0.60(0.47-0.76) | <0.001 |
| Xuebijing | 2.56(1.93-3.39) | <0.001 |
| Diammonium glycyrrhetate | 0.52(0.37-0.73) | <0.001 |
| Methylprednisolone | 12.07(8.99-16.22) | <0.001 |
| Dexamethasone | 2.60(2.01-3.35) | <0.001 |
| Interferon-alpha | 0.98(0.77-1.26) | 0.883 |
| Abbreviations: HR: hazard ratio; 95%CI: confidence interval | | |

**Table S3. Baseline characteristics of patients by Qingfei Paidu Tang use**

|  | **No QPT**  N=6 106 | **QPT**  N=2 833 | **SMD before IPTW** | **SMD after IPTW** |
| --- | --- | --- | --- | --- |
| **Demographic** |  |  |  |  |
| Women | 3260 (53.4%) | 1511 (53.3%) | -0.0011 | 0.0134 |
| Age, years |  |  | 0.1244 | 0.0196 |
| < 60 | 3450 (56.5%) | 1770 (62.5%) |  |  |
| 60-70 | 1461 (23.9%) | 605 (21.4%) |  |  |
| > 70 | 1195 (19.6%) | 458 (16.2%) |  |  |
| **Prior history/Comorbidities** |  |  |  |  |
| Hypertension | 2107 (34.5%) | 909 (32.1%) | -0.0514 | 0.0163 |
| Diabetes | 979 (16.0%) | 378 (13.3%) | -0.0761 | 0.0020 |
| Coronary heart disease | 458 (7.5%) | 228 (8.0%) | 0.0204 | 0.0243 |
| Stroke | 447 (7.3%) | 162 (5.7%) | -0.0649 | 0.0541 |
| Chronic kidney disease | 153 (2.5%) | 63 (2.2%) | -0.0186 | 0.0230 |
| COPD | 114 (1.9%) | 43 (1.5%) | -0.0271 | 0.0167 |
| Cancer | 192 (3.1%) | 93 (3.3%) | 0.0078 | 0.0103 |
| **Clinical characteristics at admission** |  |  |  |  |
| SBP, median (IQR), mmHg | 130(120, 140) | 128(120, 140) | -0.0240 | 0.0357 |
| DBP, median (IQR), mmHg | 80(74, 88) | 80(74, 89) | 0.0486 | 0.0279 |
| HR, median (IQR), breaths per min | 84(78, 95) | 84(78, 96) | 0.0392 | 0.0239 |
| RR >24 breaths per min | 578 (9.5%) | 209 (7.4%) | -0.0753 | 0.0598 |
| Critical state at admission | 268 (4.4%) | 122 (4.3%) | -0.0041 | 0.0079 |
| **Medication** |  |  |  |  |
| **Antiviral** |  |  |  |  |
| Arbidol | 3307 (54.2%) | 2109 (74.4%) | 0.4332 | 0.2892 |
| Ribavirin | 1068 (17.5%) | 667 (23.5%) | 0.1503 | -0.0770 |
| Oseltamivir | 1302 (21.3%) | 711 (25.1%) | 0.0895 | -0.0808 |
| Ganciclovir | 312 (5.1%) | 194 (6.8%) | 0.0734 | -0.1256 |
| Lopinavir/Ritonavir | 746 (12.2%) | 402 (14.2%) | 0.0583 | -0.0884 |
| **Traditional Chinese medicine** |  |  |  |  |
| Lianhua Qingwen | 3069 (50.3%) | 1666 (58.8%) | 0.1722 | 0.0649 |
| Xuebijing | 536 (8.8%) | 591 (20.9%) | 0.3451 | 0.0291 |
| Diammonium glycyrrhetate | 964 (15.8%) | 347 (12.2%) | -0.1021 | 0.0053 |
| **Corticosteroids** |  |  |  |  |
| Methylprednisolone | 1185 (19.4%) | 556 (19.6%) | 0.0055 | -0.1326 |
| Dexamethasone | 313 (5.1%) | 154 (5.4%) | 0.0139 | -0.0060 |
| **Immunomodulator** |  |  |  |  |
| Interferon-alpha | 2117 (34.7%) | 982 (34.7%) | -0.0002 | -0.1982 |
| Abbreviations: QPT, Qingfei Paidu Tang; IPTW, inverse probability of treatment weighting; SMD, standardized mean difference; IQR, inter-quartile range; SBP, systolic blood pressure; DBP, diastolic blood pressure; HR, heart rate; RR, respiratory rate; COPD, chronic obstructive pulmonary disease. | | | | |

**Table S4. Association between Qingfei Paidu Tang use and mortality, acute hepatic injury, acute kidney injury**

| **Analysis** | **Outcomes** |
| --- | --- |
|  | **Mortality** |
| No. of events/no. of patients at risk (%) |  |
| Qingfei Paidu Tang | 39(1.4) |
| No Qingfei Paidu Tang | 295(4.8) |
| Propensity-score analysis with IPTW, HR (95% CI) | 0.47 (0.36-0.62) |
|  | **Acute hepatic injury** |
| No. of events/no. of patients at risk (%) |  |
| Qingfei Paidu Tang | 253(8.9) |
| No Qingfei Paidu Tang | 609(10.0) |
| Propensity-score analysis with IPTW, OR (95% CI) | 0.96 (0.81-1.14) |
|  | **Acute kidney injury** |
| No. of events/no. of patients at risk (%) |  |
| Qingfei Paidu Tang | 48(1.7) |
| No Qingfei Paidu Tang | 186(3.0) |
| Propensity-score analysis with IPTW, OR (95% CI) | 0.80(0.59-1.09) |
| Abbreviations: IPTW, inverse probability of treatment weighting; HR, hazard ratio; OR, odds ratio; 95% CI: 95% confidence interval | |

**Table S5. Baseline characteristics of patients by Qingfei Paidu Tang use with propensity score matching**

|  | **No QPT** | **QPT** | **SMD** |
| --- | --- | --- | --- |
|  | N=1746 | N=1746 |  |
| **Demographic** |  |  |  |
| Women | 940 (50.8%) | 921 (52.7%) | -0.0241 |
| Age, years |  |  | 0.0297 |
| < 60 | 1071 (61.3%) | 1046 (59.9%) |  |
| 60-70 | 392 (22.5%) | 409 (23.4%) |  |
| > 70 | 283 (16.2%) | 291 (16.7%) |  |
| **Prior history/Comorbidities** |  |  |  |
| Hypertension | 526 (30.1%) | 559 (32.0%) | 0.0408 |
| Diabetes | 204 (11.7%) | 228 (13.1%) | 0.0418 |
| Coronary heart disease | 78 (4.5%) | 106 (6.1%) | 0.0718 |
| Stroke | 73 (4.2%) | 73 (4.2%) | <0.0001 |
| Chronic kidney disease | 24 (1.4%) | 22 (1.3%) | -0.0100 |
| COPD | 20 (1.1%) | 19 (1.1%) | -0.0055 |
| Cancer | 25 (1.4%) | 33 (1.9%) | 0.0359 |
| **Clinical characteristics at admission** |  |  |  |
| SBP, mmHg (IQR), mmHg | 130(120, 140) | 128(120, 140) | -0.0302 |
| DBP, median (IQR), mmHg | 80(74, 89) | 80(75, 88) | -0.0282 |
| HR, median (IQR), breaths per min | 84(78, 96) | 84(78, 95) | -0.0119 |
| RR >24 breaths per min | 121 (6.9%) | 136 (7.8%) | 0.0329 |
| Critical state at admission | 44 (2.5%) | 50 (2.9%) | 0.0212 |
| **Medication** |  |  |  |
| **Antiviral** |  |  |  |
| Arbidol | 961 (55.0%) | 1248 (71.5%) | 0.3460 |
| Ribavirin | 328 (18.8%) | 263 (15.1%) | -0.0994 |
| Oseltamivir | 318 (18.2%) | 348 (19.9%) | 0.0437 |
| Ganciclovir | 74 (4.2%) | 41 (2.3%) | -0.1061 |
| Lopinavir/Ritonavir | 183 (10.5%) | 148 (8.5%) | -0.0685 |
| **Traditional Chinese medicine** |  |  |  |
| Lianhua Qingwen | 849 (48.6%) | 1033 (59.2%) | 0.2126 |
| Xuebijing | 199 (11.4%) | 175 (10.0%) | -0.0445 |
| Diammonium glycyrrhetate | 241 (13.8%) | 217 (12.4%) | -0.0407 |
| **Corticosteroids** |  |  |  |
| Methylprednisolone | 262 (15.0%) | 199 (11.4%) | -0.1067 |
| Dexamethasone | 90 (5.2%) | 71 (4.1%]) | -0.0519 |
| **Immunomodulator** |  |  |  |
| Interferon-alpha | 546 (31.3%) | 384 (22.0%) | -0.2111 |
| Abbreviations: QPT, Qingfei Paidu Tang; IPTW, inverse probability of treatment weighting; SMD, standardized mean difference; IQR, inter-quartile range; HR, heart rate; RR, respiratory rate; COPD: chronic obstructive pulmonary disease | | | |
